# BioBERT-Derived Clinical Text Representations and Sparse Principal Components of Multi-Omics Data for Breast Cancer Survival Prediction: A Leakage-Controlled Cross-Validated Benchmark

**DOI:** 10.64898/2026.08.31.26361875

**Authors:** Md Mehedi Hasan Bhuiyan

## Abstract

**Background:** Reported performance for multi-modal cancer prognostic models is frequently obtained from a single train–test partition and without comparison against a simple clinical baseline. We evaluated an integrated framework combining language-model representations of structured clinical records with high-dimensional multi-omics data under a leakage-controlled protocol.

**Methods:** Clinical variables from 589 TCGA-BRCA patients were converted to natural language sentences and encoded with BioBERT; DNA methylation, RNA, miRNA and protein data were reduced by Sparse principal component analysis. Clinical-only, multi-omics-only and integrated feature sets were evaluated with Cox proportional hazards, XGBoost and random survival forest (RSF) using five repetitions of stratified five-fold cross-validation with pooled out-of-fold predictions. Models were compared against a marginal Kaplan–Meier null and a conventional age-and-stage Cox model, with differences assessed by paired bootstrap.

**Results:** Among 589 patients with 88 deaths (14.9%; median follow-up 33.7 months), the integrated RSF model performed best (Harrell’s C 0.685, 95% CI 0.620–0.754; Uno’s C 0.748), exceeding the conventional clinical Cox model (0.637, 0.559–0.707). The ordering integrated > clinical-only > multi-omics-only held in all nine feature-set by algorithm comparisons and at all three component settings tested. The integrated model significantly outperformed every multi-omics-only model (*P* = 0.008–0.044) but not clinical-only models (ΔC = 0.037, *P* = 0.128). Brier scores approached a null reference at 2 and 3 years and improved modestly at 5 years.

**Conclusion:** Integration produced the highest discrimination and exceeded a conventional clinical model, but the gain over clinical features within the same pipeline was not statistically resolvable at this event count. Reporting simple baselines, null-model calibration and cross-validated estimates should be standard in multi-modal prognostic modelling.

## 1 Introduction

Breast cancer is among the most frequently diagnosed malignancies worldwide and exhibits substantial heterogeneity in clinical presentation, molecular characteristics, therapeutic response and survival outcomes [1]. Accurate survival prediction underpins personalised oncology. Conventional prognostic models rely on demographic and clinicopathologic variables such as age, tumour stage, histologic subtype and treatment [2, 3]. These variables carry substantial prognostic information but do not directly capture the molecular processes driving tumour progression.

High-throughput sequencing enables simultaneous measurement of multiple molecular layers—DNA methylation, mRNA expression, microRNA expression and protein expression— creating opportunities for multi-omics survival modelling [4–6]. Each layer characterises distinct biology: methylation reflects epigenetic alteration, RNA captures transcriptional activity, miRNA governs post-transcriptional regulation and protein expression reflects functional cellular state [5, 7]. These data are, however, extremely high-dimensional relative to available sample sizes (*N* ≪ *P*), which raises a substantial risk of overfitting and optimistic performance estimates.

Recent large-scale evaluations have tempered expectations for multi-omics survival modelling. In a benchmark across multiple cancer cohorts, Herrmann et al. found that only one of several multi-omics methods outperformed a Cox model on average, and that for six of the datasets examined the Cox model was not outperformed at all [8]. The standardised SurvBoard benchmark similarly reported that regularised linear and random-forest methods matched or exceeded deep learning approaches, and that multi-omics did not improve on a clinical-only baseline in TCGA [9]. A recurring feature of studies reporting stronger results is evaluation on a single train–test partition, often with feature selection or dimensionality reduction estimated before partitioning.

Dimensionality reduction is therefore essential prior to survival modelling [10, 11]. Sparse Principal Component Analysis extends standard PCA with an *L*_1_ penalty on the loading vectors, producing sparse components that retain influential feature subsets [10]. Because Sparse PCA is unsupervised, the extracted components capture directions of maximal variance rather than of maximal prognostic association—a trade-off we examine explicitly.

In parallel, clinical characteristics can be represented using natural language processing. Transformer architectures capture bidirectional semantic relationships through self-attention [12, 13], and domain-specific models such as BioBERT, pretrained on biomedical literature, provide contextual understanding of clinical terminology [14, 15]. Sentence-BERT frameworks yield fixed-size semantic representations [16]. Converting structured records into patient-level narratives and encoding them with a biomedical language model maps heterogeneous categorical and continuous factors into a unified latent space. Whether this encoding preserves the information carried by the source variables is, however, an empirical question that requires direct comparison against those variables—a comparison we report here.

We evaluated a survival prediction framework coupling BioBERT-derived clinical embeddings with Sparse PCA representations of four molecular modalities, under repeated cross-validation with pooled out-of-fold predictions. We benchmarked clinical-only, multiomics-only and integrated feature sets across regularised CoxPH, XGBoost and RSF [17–19], and compared every model against both a marginal Kaplan–Meier reference and conventional Cox models fitted directly to the structured clinical variables.

## 2 Materials and Methods

### 2.1 Data Source and Cohort Construction

Breast cancer data were obtained from the UCSC Xena platform, which curates harmonised clinical, genomic and molecular datasets from TCGA [7]. The TCGA-BRCA cohort provided matched demographic, clinicopathologic, DNA methylation, RNA expression, miRNA expression and RPPA protein expression data, aligned by patient identifier. Patients lacking matched multi-omics profiles, missing essential clinical annotation, or with non-positive overall survival time (OS.time ≤ 0) were excluded, yielding 589 patients. The analytical workflow is shown in Figure 1.

**Figure 1:**
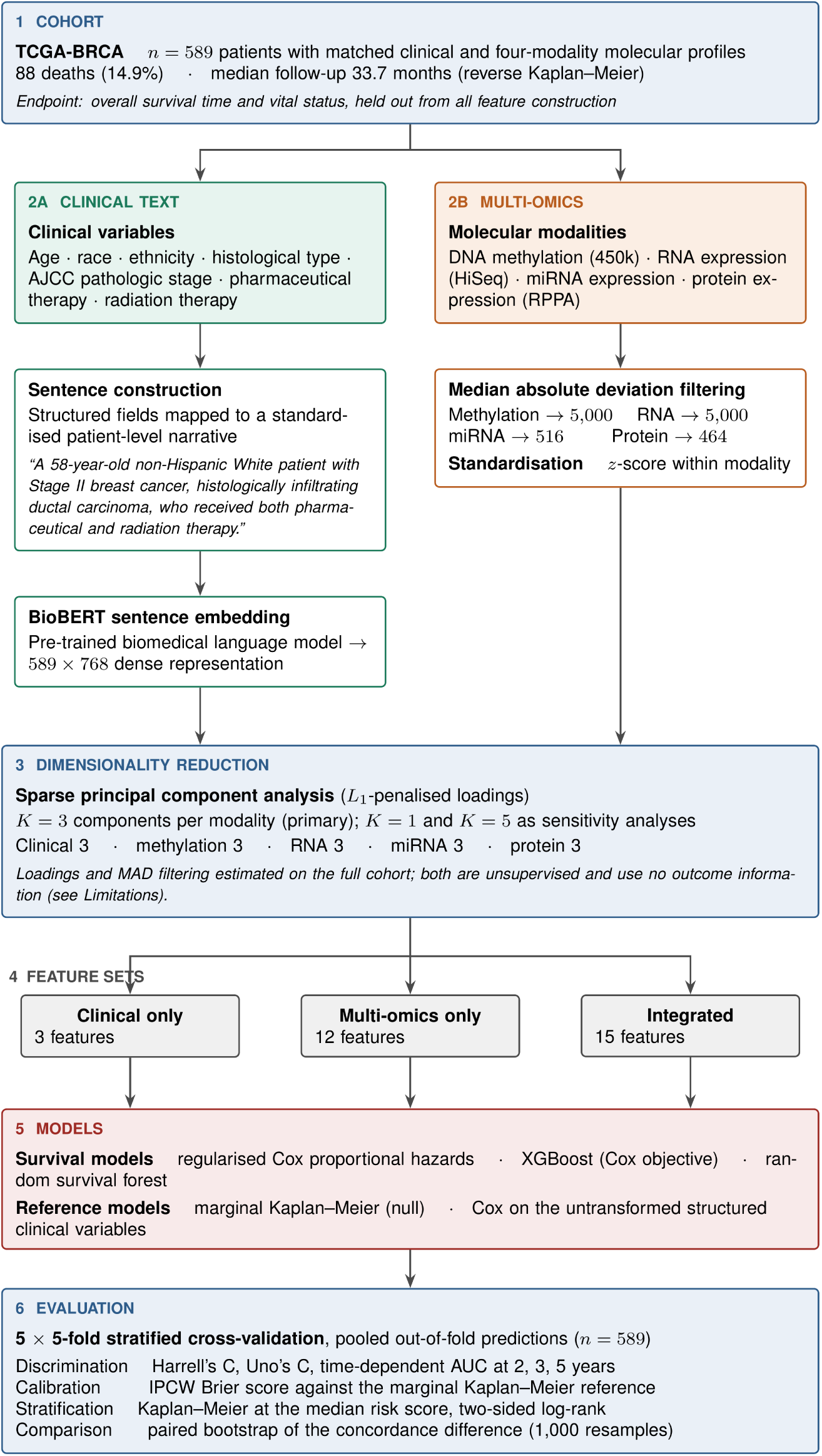
Analytical workflow: cohort construction, conversion of structured clinical variables to natural language sentences, BioBERT sentence embedding, median absolute deviation filtering and Sparse PCA of the molecular modalities, and cross-validated survival modelling.

### 2.2 Clinical Variables and Sentence Construction

The clinical predictor set comprised age at diagnosis, race, ethnicity, disease histological type, AJCC pathologic stage, pharmaceutical therapy status and radiation therapy status. Overall survival time and vital status were reserved as outcome variables and excluded from feature construction. Structured variables were mapped to standardised patient-level natural language sentences integrating demographic profile, tumour pathology and adjuvant treatment history. Survival duration and vital status were excluded from text synthesis.

Adjuvant therapy status is recorded in TCGA as an ordered pair of treatment types with a corresponding pair of administration indicators. The ordering of this pair varies between records; indicators were therefore matched to treatment type by name rather than by position. Pathologic stage was collapsed to Stage I–IV, because several of the eleven recorded sub-stages contained fewer than five patients.

### 2.3 Multi-Omics Processing and BioBERT Embedding

Molecular datasets comprised DNA methylation (Illumina 450k), RNA expression (Illumina HiSeq RNASeqV2), miRNA expression and protein expression (RPPA). Median absolute deviation filtering isolated the most variable features within each layer, producing matrices of dimension 589 × 5000 (methylation), 589 × 5000 (RNA), 589 × 516 (miRNA) and 589 × 464 (protein). Clinical narratives were tokenised and passed through a BioBERT-based SentenceTransformer model [13, 14], producing a 768-dimensional embedding per patient.

### 2.4 Sparse Principal Component Analysis

Sparse PCA [10] was applied independently to the clinical embeddings and to each molecular matrix. The primary analysis retained *K* = 3 components per modality, giving feature sets of dimension 3 (clinical-only), 12 (multi-omics-only) and 15 (integrated). Settings *K* = 1 and *K* = 5 were evaluated as sensitivity analyses.

Sparse PCA loadings and median absolute deviation filtering were estimated on the full cohort prior to partitioning. Both procedures are unsupervised and do not use survival time or vital status; nonetheless, this represents a deviation from strict within-fold preprocessing and is noted as a limitation. All supervised model fitting, hyperparameter selection and risk-threshold derivation were performed within training folds only.

### 2.5 Evaluation Design

Performance was estimated by five repetitions of stratified five-fold cross-validation. Folds were stratified on event status crossed with follow-up tertile so that each fold carried a comparable censoring pattern. Within each repetition every patient contributed exactly one out-of-fold prediction; predictions were pooled across folds, so that discrimination, calibration and Kaplan–Meier analyses used all 589 patients while no patient’s own data contributed to their predicted risk. A conventional single 70/30 partition is reported in Supplementary Table S2 for comparison. All reference models were fitted using the identical folds, seed and stratification, so their concordance indices are directly comparable.

### 2.6 Survival Models

1. **Regularised CoxPH:** *L*_2_ ridge penalty (*α* = 0.1) [17].
2. **XGBoost (survival:cox):** gradient-boosted trees with the Cox partial likelihood objective (learning rate 0.05, maximum depth 3, 100 boosting rounds) [18].
3. **Random survival forest:** log-rank splitting, 300 trees, minimum leaf size 5 [19], implemented in scikit-survival.
4. **Marginal Kaplan–Meier reference:** the training-fold survival curve applied to every test patient, providing a null model against which calibration is assessed.
5. **Conventional clinical Cox:** a Cox model fitted to age (standardised) and grouped AJCC stage, the standard prognostic baseline in breast cancer. Two extended models— additionally including race, and additionally including adjuvant therapy—are reported in Supplementary Table S4. Adjuvant therapy was excluded from the primary baseline because it is assigned after diagnosis and is influenced by prognosis.

### 2.7 Performance Metrics

- **Harrell’s concordance index** [2], with 95% confidence intervals from 1,000 bootstrap resamples.
- **Uno’s concordance index** [20], an inverse-probability-of-censoring-weighted estimator that is not biased by the censoring distribution. This is reported because 85% of the cohort is censored.
- **Time-dependent cumulative/dynamic AUC** at 2, 3 and 5 years, computed with IPCW weights [21, 22]. One-year AUC was not evaluated: only 11 deaths occurred within 12 months across the entire cohort. Horizons beyond 5 years were not evaluated because fewer than 20 patients remained at risk.
- **IPCW Brier score** [23] at 2, 3 and 5 years, reported alongside the marginal Kaplan–Meier reference.
- **Kaplan–Meier stratification** at the median cross-validated risk score, with two-sided log-rank tests and number-at-risk tables.
- **Paired bootstrap comparison:** differences in concordance were assessed by resampling patients 1,000 times, re-evaluating both models on each resample, and constructing confidence intervals and two-sided *P* values from the distribution of the paired difference.

Reporting follows the TRIPOD+AI statement [24]. Analyses used Python 3.10 with scikit-learn, lifelines, scikit-survival and xgboost. Random seeds and package versions are recorded in the accompanying repository.

## 3 Results

### 3.1 Cohort Characteristics

Baseline characteristics are summarised in Table 1. The cohort comprised 589 patients with 88 deaths (14.9%) and a median follow-up of 33.7 months by reverse Kaplan–Meier estimation. Overall survival was 95.0%, 88.9% and 78.9% at 2, 3 and 5 years. Mean age was 58.1 ± 13.0 years. Patients were predominantly White (*n* = 454, 77.1%), and ductal and lobular neoplasms accounted for 95.2% of cases. Stage II was most prevalent (*n* = 368, 62.5%). Pharmaceutical and radiation therapy were recorded as administered in 79.3% and 51.1% of patients respectively, with therapy status unrecorded in 8.7%. Median overall survival was not reached within the range of follow-up that can be estimated reliably; only 16 patients remained at risk beyond 120 months.

**Table 1:**
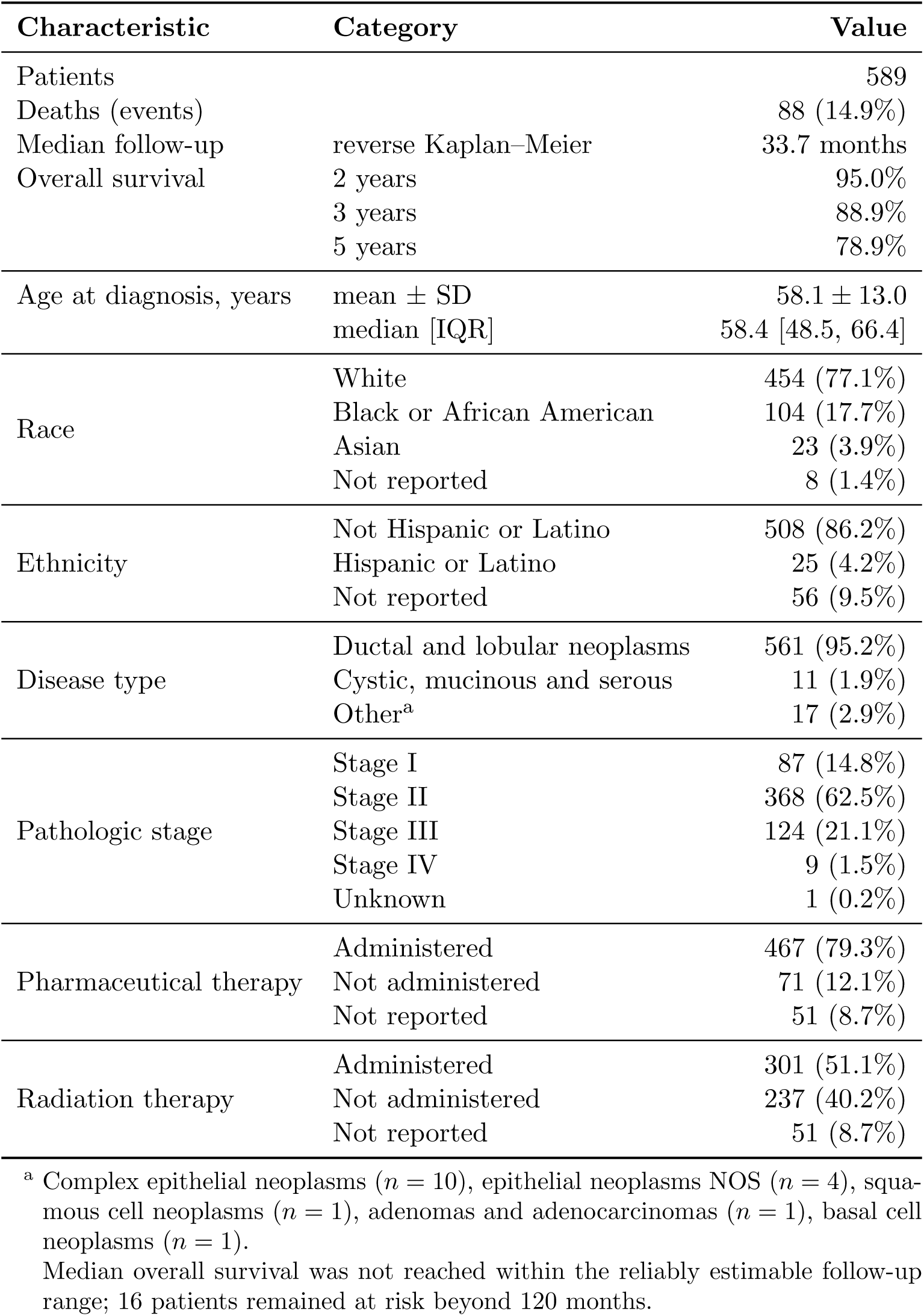
Baseline characteristics of the study cohort (*n* = 589).

| Characteristic | Category | Value |
| --- | --- | --- |
| Patients |  | 589 |
| Deaths (events) |  | 88 (14.9%) |
| Median follow-up | reverse Kaplan–Meier | 33.7 months |
| Overall survival | 2 years | 95.0% |
|  | 3 years | 88.9% |
|  | 5 years | 78.9% |
| Age at diagnosis, years | mean ± SD | 58.1 ± 13.0 |
|  | median [IQR] | 58.4 [48.5, 66.4] |
| Race | White | 454 (77.1%) |
|  | Black or African American | 104 (17.7%) |
|  | Asian | 23 (3.9%) |
|  | Not reported | 8 (1.4%) |
| Ethnicity | Not Hispanic or Latino | 508 (86.2%) |
|  | Hispanic or Latino | 25 (4.2%) |
|  | Not reported | 56 (9.5%) |
| Disease type | Ductal and lobular neoplasms | 561 (95.2%) |
|  | Cystic, mucinous and serous | 11 (1.9%) |
|  | Other<sup>a</sup> | 17 (2.9%) |
| Pathologic stage | Stage I | 87 (14.8%) |
|  | Stage II | 368 (62.5%) |
|  | Stage III | 124 (21.1%) |
|  | Stage IV | 9 (1.5%) |
|  | Unknown | 1 (0.2%) |
| Pharmaceutical therapy | Administered | 467 (79.3%) |
|  | Not administered | 71 (12.1%) |
|  | Not reported | 51 (8.7%) |
| Radiation therapy | Administered | 301 (51.1%) |
|  | Not administered | 237 (40.2%) |
|  | Not reported | 51 (8.7%) |

<sup>a</sup> Complex epithelial neoplasms ( $n = 10$ ), epithelial neoplasms NOS ( $n = 4$ ), squamous cell neoplasms ( $n = 1$ ), adenomas and adenocarcinomas ( $n = 1$ ), basal cell neoplasms ( $n = 1$ ).
Median overall survival was not reached within the reliably estimable follow-up range; 16 patients remained at risk beyond 120 months.

### 3.2 Discrimination

Cross-validated discrimination is shown in Table 2. The integrated feature set achieved the highest observed discrimination with RSF (Harrell’s C 0.685, 95% CI 0.620–0.754; Uno’s C 0.748), followed by integrated XGBoost (0.678, 0.605–0.750) and integrated CoxPH (0.658, 0.588–0.728). The best clinical-only model was RSF (0.648, 0.569–0.732) and the best multiomics-only model was RSF (0.618, 0.554–0.683). Fold-level concordance distributions are shown in Supplementary Figure S5.

**Table 2:** Cross-validated discrimination on pooled out-of-fold predictions (*K* = 3 sparse principal components per modality; five repetitions of stratified five-fold cross-validation; *n* = 589, 88 events).

| Feature set | Model | Harrell’s C<br>(95% CI) | Uno’s C | $\Delta C$ vs reference (95% CI), $P$ |
| --- | --- | --- | --- | --- |
| *Reference models* |  |  |  |  |
| Marginal Kaplan–Meier | — | 0.500 | 0.500 | — |
| Clinical Cox<sup>a</sup> | age + stage | 0.637 (0.559–0.707) | 0.549 | — |
| *Clinical only* (3 features) |  |  |  |  |
|  | CoxPH | 0.645 (0.567–0.721) | 0.693 | +0.040 (−0.018, +0.100), 0.164 |
|  | XGBoost | 0.642 (0.562–0.728) | 0.715 | +0.044 (−0.012, +0.100), 0.132 |
|  | RSF | 0.648 (0.569–0.732) | 0.719 | +0.037 (−0.012, +0.089), 0.128 |
| *Multi-omics only* (12 features) |  |  |  |  |
|  | CoxPH | 0.583 (0.512–0.656) | 0.631 | +0.103 (+0.025, +0.183), **0.008** |
|  | XGBoost | 0.606 (0.537–0.676) | 0.636 | +0.079 (+0.014, +0.153), **0.028** |
|  | RSF | 0.618 (0.554–0.683) | 0.655 | +0.068 (+0.003, +0.142), **0.044** |
| *Integrated* (15 features) |  |  |  |  |
|  | CoxPH | 0.658 (0.588–0.728) | 0.709 | +0.027 (−0.020, +0.078), 0.232 |
|  | XGBoost | 0.678 (0.605–0.750) | 0.725 | +0.008 (−0.027, +0.047), 0.628 |
|  | RSF | **0.685 (0.620–0.754)** | **0.748** | reference |

<sup>a</sup> Cox proportional hazards model fitted to standardised age and grouped AJCC stage, the conventional clinical prognostic baseline. Extended models additionally including race and adjuvant therapy are reported in Supplementary Table S4; adjuvant therapy was excluded from the primary baseline because it is assigned after diagnosis and is influenced by prognosis.
RSF, random survival forest.

The ordering integrated > clinical-only > multi-omics-only held in all nine feature-set by algorithm comparisons. In paired bootstrap tests against the integrated RSF model, all three multi-omics-only models were significantly less discriminative (ΔC = 0.068–0.103, *P* = 0.008–0.044), whereas no clinical-only model differed significantly (ΔC = 0.037–0.044, *P* = 0.128–0.164). The three algorithms applied to the integrated feature set did not differ significantly from one another (*P* = 0.232 and *P* = 0.628 for CoxPH and XGBoost versus RSF).

### 3.3 Comparison with Conventional Clinical Models

A Cox model fitted to age and grouped pathologic stage, the conventional prognostic baseline, achieved a pooled out-of-fold Harrell’s C of 0.637 (95% CI 0.559–0.707), which the integrated RSF model exceeded by 0.048. The difference was more pronounced on Uno’s C (0.748 versus 0.549). This gap between the two concordance measures is informative: the clinical model achieved its highest time-dependent AUC at 2 years (0.697) and its lowest at 5 years (0.610), whereas the integrated model showed the opposite trajectory (0.634 rising to 0.733). Because Uno’s estimator upweights later event times, a clinical model whose discrimination is concentrated in early follow-up is penalised accordingly. Age was the only individually significant predictor in the baseline model (hazard ratio 1.47 per standard deviation, 95% CI 1.19–1.81, *P <* 0.001); Stage III versus Stage I did not reach significance (1.62, 0.85–3.08, *P* = 0.143). Adding race did not improve fit (likelihood ratio *χ*^2^ = 1.09, *df* = 2, *P* = 0.579).

A separate question is whether the BioBERT encoding preserves the information in the variables from which the sentences were generated. The appropriate comparator here is a Cox model using all seven source variables, which reached 0.679 (0.603–0.760); the BioBERT encoding of those same variables, reduced to three sparse components, reached 0.648 (0.569– 0.732). The encoding therefore did not improve on direct use of the source variables and appears to have lost a modest amount of information. We note that this extended Cox model includes adjuvant therapy, which is assigned after diagnosis; its apparent contribution (hazard ratio 0.18 for combined therapy versus none, *P <* 0.001) most plausibly reflects confounding by indication rather than prognostic information available at diagnosis. It is reported as an information-preservation comparator, not as a prognostic baseline (Supplementary Table S4).

### 3.4 Calibration

IPCW Brier scores are shown in Table 3 alongside the marginal Kaplan–Meier reference. At 2 years the null model achieved 0.047 and seven of nine models were equal to or worse than this value; at 3 years the null achieved 0.099 and two models were worse. At 5 years the integrated RSF model achieved 0.151 against a null of 0.168, the largest improvement observed. Discrimination therefore exceeded that of the null model substantially, whereas absolute survival probability estimates improved only modestly and only at the longest horizon. Equivalent results for *K* = 1 and *K* = 5 are shown in Supplementary Figures S7 and S8.

**Table 3:** Time-dependent discrimination and calibration on pooled out-of-fold predictions (*K* = 3).

| Feature set | Model | Time-dependent AUC |  |  | IPCW Brier score |  |  |
| --- | --- | --- | --- | --- | --- | --- | --- |
|  |  | 2 y | 3 y | 5 y | 2 y | 3 y | 5 y |
| Marginal Kaplan–Meier | — | 0.500 | 0.500 | 0.500 | 0.047 | 0.099 | 0.168 |
| *Clinical only* |  |  |  |  |  |  |  |
|  | CoxPH | 0.617 | 0.543 | 0.637 | 0.047 | 0.099 | 0.157 |
|  | XGBoost | 0.576 | 0.585 | 0.697 | 0.049 | 0.105 | 0.154 |
|  | RSF | 0.573 | 0.604 | 0.703 | 0.048 | 0.099 | 0.149 |
| *Multi-omics only* |  |  |  |  |  |  |  |
|  | CoxPH | 0.559 | 0.644 | 0.598 | 0.048 | 0.097 | 0.165 |
|  | XGBoost | 0.557 | 0.656 | 0.610 | 0.049 | 0.098 | 0.178 |
|  | RSF | 0.568 | **0.688** | 0.637 | 0.048 | 0.096 | 0.165 |
| *Integrated* |  |  |  |  |  |  |  |
|  | CoxPH | 0.634 | 0.628 | 0.671 | 0.048 | 0.098 | 0.159 |
|  | XGBoost | 0.624 | 0.654 | 0.709 | 0.049 | 0.103 | 0.171 |
|  | RSF | **0.634** | 0.668 | **0.733** | **0.047** | **0.097** | **0.151** |

Estimates are based on 24, 43 and 64 deaths at 2, 3 and 5 years respectively. One-year values are not reported because only 11 deaths occurred within 12 months across the cohort.
IPCW, inverse probability of censoring weighting; RSF, random survival forest.

### 3.5 Time-Dependent Discrimination

Time-dependent AUC for the integrated RSF model was 0.634, 0.668 and 0.733 at 2, 3 and 5 years (Figure 3), based on 24, 43 and 64 deaths respectively. Multi-omics-only features discriminated most strongly at 3 years (AUC 0.688 for RSF) and least strongly at 2 years (0.568). Clinical-only features under the Cox model showed the opposite ordering, with a higher 2-year than 3-year AUC (0.617 versus 0.543), although this pattern was not reproduced by the two ensemble methods and should be interpreted cautiously given that the 2-year estimate rests on 24 deaths.

### 3.6 Risk Stratification

Stratification at the median cross-validated risk score separated overall survival for every feature set and algorithm (Table 4, Figure 2). Log-rank *χ*^2^ statistics followed the same ordering as the concordance results: integrated (23.9–30.6) > clinical-only (14.7–20.6) > multi-omics-only (10.7–16.9). For the integrated RSF model, 24 deaths occurred among 294 low-risk patients compared with 64 among 295 high-risk patients (*χ*^2^ = 30.6, *P <* 0.001).

**Figure 2:**
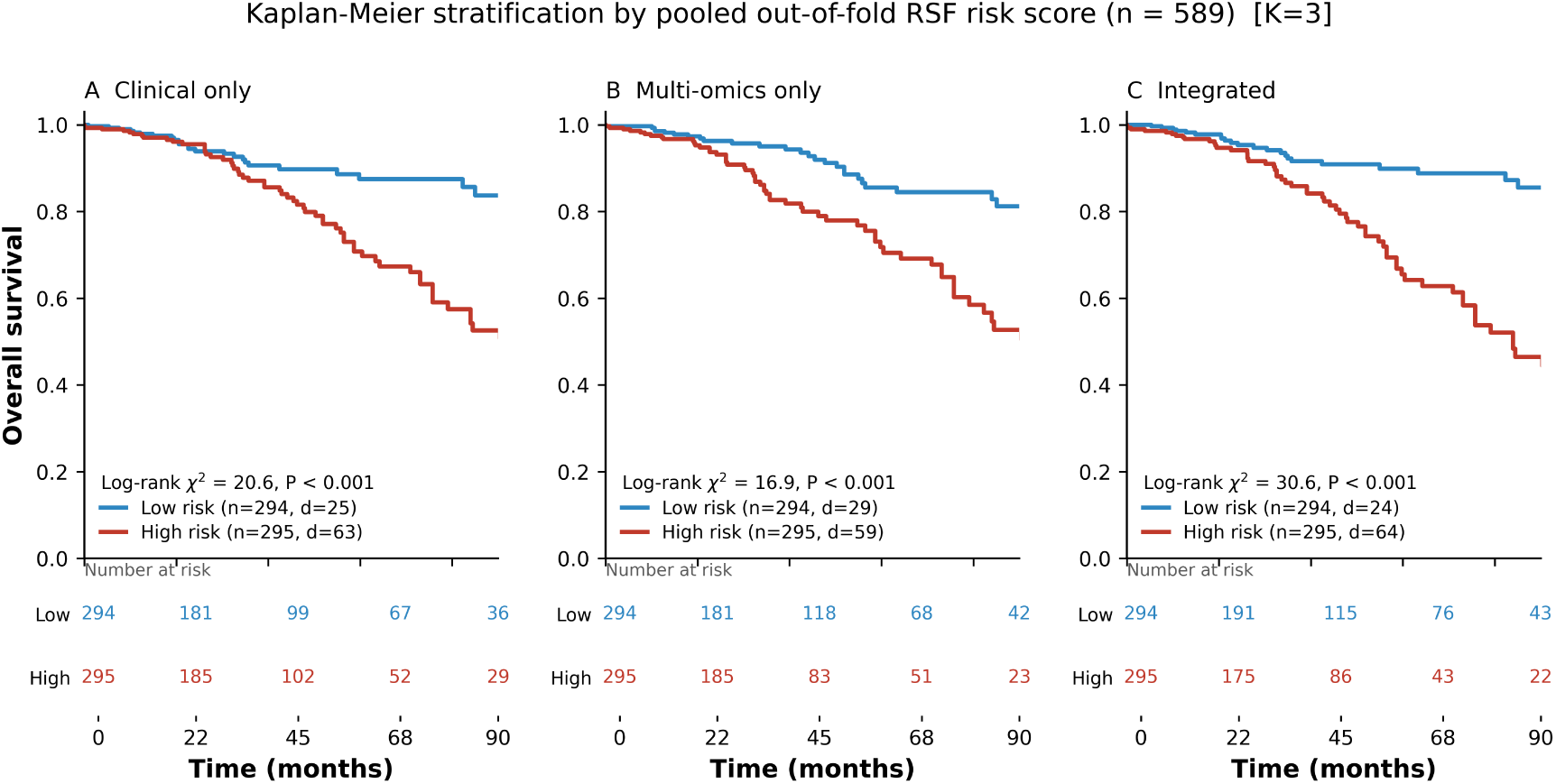
Kaplan–Meier overall survival by predicted risk group for random survival forest models fitted to clinical-only (A), multi-omics-only (B) and integrated (C) feature sets (*K* = 3). Patients were dichotomised at the median cross-validated risk score. Risk scores are pooled out-of-fold predictions from five repetitions of stratified five-fold cross-validation across all 589 patients, so no patient’s own data contributed to their predicted risk. Curves are truncated at 90 months, beyond which fewer than 10% of patients remain at risk. *d* denotes deaths; *P* values are two-sided log-rank tests.

**Figure 3:**
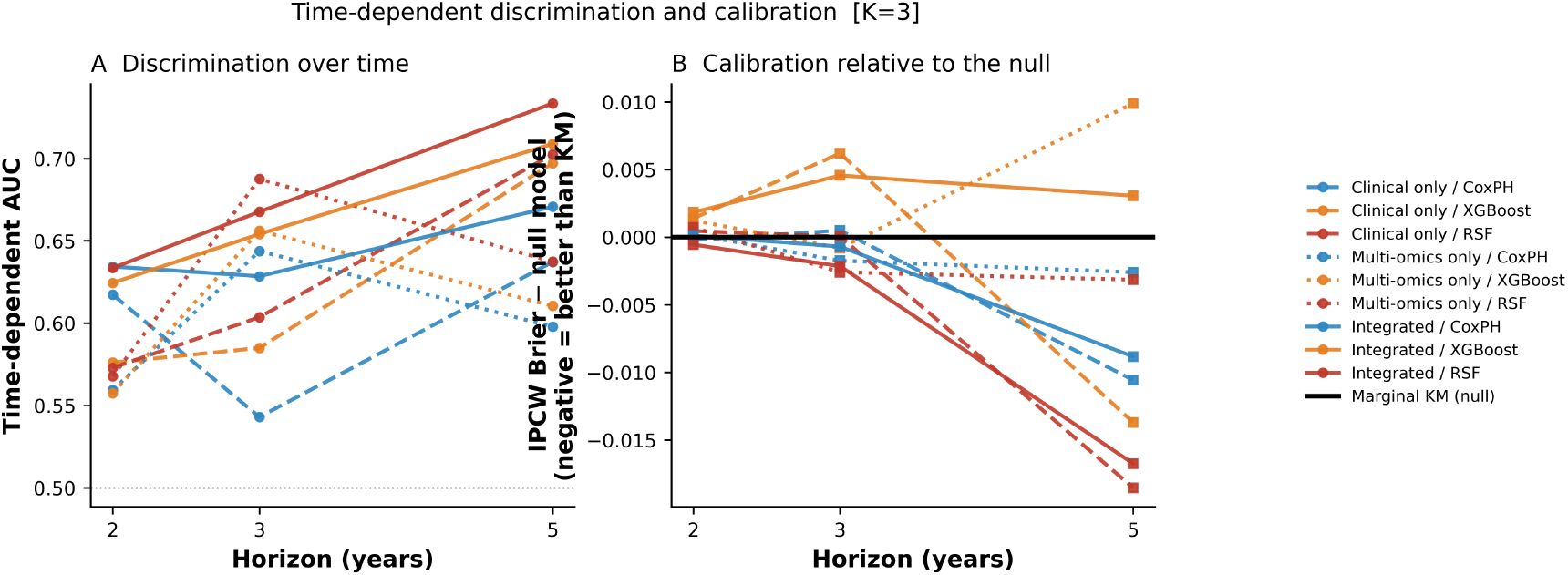
Time-dependent discrimination and calibration from pooled out-of-fold predictions (*K* = 3). (A) Cumulative/dynamic AUC at 2, 3 and 5 years, based on 24, 43 and 64 deaths respectively. (B) IPCW Brier score expressed as a difference from a marginal Kaplan–Meier reference; values below zero indicate better calibration than the population survival curve.

**Table 4:** Kaplan–Meier risk stratification at the median cross-validated risk score (*K* = 3, pooled out-of-fold predictions, *n* = 589).

| Feature set | Model | Log-rank $\chi^2$ | $P$ | Deaths, low risk | Deaths, high risk |
| --- | --- | --- | --- | --- | --- |
| Clinical only | CoxPH | 14.7 | $< 0.001$ | 27/294 | 61/295 |
| | XGBoost | 15.8 | $< 0.001$ | 28/294 | 60/295 |
| | RSF | 20.6 | $< 0.001$ | 25/294 | 63/295 |
| Multi-omics only | CoxPH | 11.0 | $< 0.001$ | 28/294 | 60/295 |
|  | XGBoost | 10.7 | 0.001 | 30/294 | 58/295 |
| | RSF | 16.9 | $< 0.001$ | 29/294 | 59/295 |
| Integrated | CoxPH | 23.9 | $< 0.001$ | 23/294 | 65/295 |
| | XGBoost | 26.7 | $< 0.001$ | 22/294 | 66/295 |
|  | RSF | **30.6** | **$< 0.001$** | 24/294 | 64/295 |

All stratifications reached statistical significance. This differs from earlier analyses of these data based on a 177-patient held-out set and reflects the larger effective sample available from pooled out-of-fold prediction rather than improved model performance; comparisons between feature sets should therefore be based on the magnitude of the *χ*^2^ statistic rather than on significance thresholds. Corresponding Kaplan–Meier curves for all nine feature-set by algorithm combinations are given in Supplementary Figure S2.

### 3.7 Sensitivity to the Number of Sparse Components

Results for *K* = 1, 3 and 5 components per modality are given in Supplementary Table S1. Integrated discrimination was essentially unchanged across settings (best C 0.685, 0.685 and 0.711 respectively, with overlapping confidence intervals). Multi-omics-only discrimination was lowest at *K* = 1 (0.572, 0.505–0.637), highest at *K* = 3 (0.618, 0.554–0.683) and intermediate at *K* = 5 (0.604, 0.533–0.670), indicating that a single component per modality under-represents the molecular layers. Kaplan–Meier stratification and cross-validated concordance distributions for each component setting are shown in Supplementary Figures S1–S6, and time-dependent discrimination across settings in Supplementary Figures S7–S9. At *K* = 5 the integrated feature set comprised 25 variables against 88 events (3.5 events per variable), below conventional recommendations [25]; *K* = 3 was therefore selected for the primary analysis.

The number of components was not pre-specified. The selection criteria stated above were applied after the sensitivity analysis was performed, and all three settings are reported in full in Supplementary Tables S1 and S3.

### 3.8 Comparison with the Single-Split Analysis

Under a conventional single 70/30 partition (Supplementary Table S2), the integrated RSF model achieved C = 0.669 (0.545–0.803), with time-dependent AUC based on 7, 15 and 20 deaths at 2, 3 and 5 years in the 177-patient test set. Confidence intervals were approximately twice as wide as those obtained by cross-validation, illustrating the limited precision available from a single partition at this event count.

## 4 Discussion

This study evaluated a multimodal survival prediction framework combining BioBERT representations of clinical narratives with Sparse PCA representations of four molecular layers, under repeated cross-validation with pooled out-of-fold predictions. Four findings emerge.

First, the integrated representation achieved the highest observed discrimination, and the ordering integrated > clinical-only > multi-omics-only was reproduced in all nine feature-set by algorithm comparisons and at all three Sparse PCA settings examined. This consistency is stronger evidence of a genuine ordering than any individual comparison, since the estimates share the same cohort but differ in model class and dimensionality.

Second, the integrated model exceeded a conventional age-and-stage Cox model by 0.048 in Harrell’s C (0.685 versus 0.637) and by a substantially larger margin in Uno’s C (0.748 versus 0.549). The divergence between the two concordance measures reflects a difference in the temporal profile of discrimination: age and stage predicted early mortality well (2-year AUC 0.697) but discriminated poorly among longer-term survivors (5-year AUC 0.610), whereas the integrated model improved steadily with horizon (0.634 to 0.733). Because Uno’s estimator upweights later event times under heavy censoring, this difference is substantial for any application concerned with medium-term prognosis. The clinical model also separated risk groups far more weakly (*χ*^2^ = 6.3 versus 30.6).

Third, the advantage over clinical features within the same pipeline was not statistically resolvable. The paired difference between the integrated and best clinical-only model was 0.037 (95% CI −0.012 to +0.089, *P* = 0.128). With 88 observed events, this cohort is underpowered to establish differences of this magnitude, and this limitation applies to any comparison of similar size reported in cohorts of this scale. By contrast, the integrated representation significantly outperformed every multi-omics-only model, indicating that the clinical component carries the majority of the prognostic signal.

Fourth, the BioBERT encoding did not improve on direct use of the variables from which the sentences were generated. A Cox model on those seven variables reached 0.679, whereas their BioBERT encoding reduced to three sparse components reached 0.648. Because the sentences are generated deterministically from the structured fields, the embedding cannot contain information those fields lack; it can only re-encode them, and in this cohort the encoding followed by unsupervised sparse dimensionality reduction lost a modest amount of information. This does not imply that language-model representations of clinical text are without value—in settings with genuinely unstructured narrative text, free-text pathology reports or clinical notes, such representations may capture information unavailable in structured fields—but it does indicate that converting already-structured variables into sentences and re-encoding them provided no measurable benefit here. Whether supervised rather than unsupervised dimensionality reduction would preserve more of the embedded information is a natural question for future work.

These findings sit within the recent benchmark literature. Herrmann et al. reported that for six of the cancer datasets they examined, multi-omics methods did not outperform a Cox model [8], and the SurvBoard benchmark found that multi-omics did not improve on a clinical-only baseline in TCGA and that statistical models generally matched or exceeded deep learning approaches, particularly in calibration [9]. Our multi-omics-only concordance of 0.618 reproduces the first of these observations closely; our integrated result is more favourable than the second, though the difference from clinical features alone was not statistically resolvable.

The calibration results deserve emphasis. Although discrimination was well above chance, IPCW Brier scores were comparable to a marginal Kaplan–Meier reference at 2 and 3 years, and better only at 5 years and only modestly (0.151 versus 0.168). Discrimination and calibration are distinct properties, and a model that ranks patients correctly may still estimate absolute risk no better than the population average. Studies reporting only concordance may therefore overstate clinical utility.

Among algorithms, random survival forest achieved the highest point estimates in most cells, but the differences between RSF, XGBoost and regularised Cox on the integrated feature set were not significant. We therefore do not conclude that ensemble methods are superior in this setting; with 15 input features the scope for higher-order interaction is limited, and the observed differences are consistent with sampling variability.

## Limitations

Several limitations should be noted. First, Sparse PCA loadings and median absolute deviation filtering were estimated on the full cohort before partitioning. Both procedures are unsupervised and do not use outcome information, so the resulting optimism is expected to be small, but this is a deviation from strict within-fold preprocessing and may contribute modest bias. Second, with 88 events, the cohort cannot resolve concordance differences below approximately 0.05; several of our comparisons fall below this threshold. Third, median follow-up of 33.7 months restricts inference about long-term survival, and horizons beyond 5 years could not be evaluated. Fourth, adjuvant therapy variables are assigned after diagnosis and are influenced by prognosis; they were therefore excluded from the primary clinical baseline, and the extended model containing them is reported only as an information-preservation comparator. Fifth, the proportional hazards assumption was not formally assessed. Sixth, clinical features were restricted to variables available in TCGA; performance status, longitudinal treatment detail and recurrence dates were unavailable. Finally, the analysis used a single retrospective cohort; external validation in an independent multi-centre cohort, such as METABRIC, is required before any claim of generalisability.

## 5 Conclusion

Under leakage-controlled repeated cross-validation, integrating BioBERT-derived clinical embeddings with Sparse PCA multi-omics features achieved the highest observed discrimination among the representations evaluated, exceeded a conventional age-and-stage Cox model, and separated survival risk groups more strongly than either source alone. The advantage over clinical features within the same pipeline was not statistically resolvable in this cohort, and the language-model encoding did not improve on direct use of the structured variables from which the sentences were generated. Molecular features alone were consistently the weakest representation. These results indicate that reporting simple clinical baselines, calibration against a null model, and cross-validated rather than single-split estimates should be standard practice in multi-modal prognostic modelling.

## Supplementary Material

Additional sensitivity analyses, cross-validated model comparisons, Kaplan–Meier analyses, and supplementary figures are provided in the **Supplementary Material**.

## Ethics Approval and Consent to Participate

This study analysed de-identified, publicly available data from The Cancer Genome Atlas obtained through the UCSC Xena platform. No institutional review board approval or informed consent was required under the TCGA data use policy for open-access data.

## Data Availability

All data analysed are publicly available from the UCSC Xena platform (https://xenabrowser.net/) under the TCGA-BRCA cohort. Processed feature matrices and patient identifiers are provided as Supplementary Data S1.

## Code Availability

All analysis code, including preprocessing, sentence construction, embedding generation, model fitting and evaluation, is available at https://github.com//mmhb100/Integrated-multiomics. Random seeds and package versions are recorded in the repository to permit exact reproduction.

## Author Contributions

M.M.H. Bhuiyan conceived the study, performed the analysis and wrote the manuscript.

## Supplementary Material

**Table S1:** Sparse PCA component sensitivity analysis. Cross-validated performance for all 27 configurations (*K* = 1, 3, 5 components per modality × three feature sets × three algorithms), pooled out-of-fold predictions, *n* = 589, 88 events.

| K | Feature set | Model | CV C-index | Pooled C (95% CI) | Uno's C | Time-dependent AUC |  |  | IPCW Brier score |  |  |
| --- | --- | --- | --- | --- | --- | --- | --- | --- | --- | --- | --- |
|  |  |  |  |  |  | 2 y | 3 y | 5 y | 2 y | 3 y | 5 y |
| K = 1 | Clinical only | CoxPH | 0.616 ± 0.082 | 0.611 (0.527–0.686) | 0.666 | 0.608 | 0.507 | 0.604 | 0.047 | 0.100 | 0.157 |
|  |  | XGBoost | 0.655 ± 0.060 | 0.638 (0.569–0.709) | 0.723 | 0.589 | 0.629 | 0.676 | 0.049 | 0.103 | 0.157 |
|  |  | RSF | 0.662 ± 0.064 | 0.662 (0.595–0.729) | 0.731 | 0.587 | 0.610 | 0.672 | 0.053 | 0.112 | 0.176 |
|  | Multi-omics only | CoxPH | 0.551 ± 0.076 | 0.553 (0.480–0.621) | 0.559 | 0.569 | 0.587 | 0.583 | 0.047 | 0.098 | 0.166 |
|  |  | XGBoost | 0.556 ± 0.075 | 0.559 (0.482–0.634) | 0.594 | 0.517 | 0.612 | 0.631 | 0.049 | 0.098 | 0.163 |
|  |  | RSF | 0.570 ± 0.078 | 0.572 (0.505–0.637) | 0.609 | 0.526 | 0.600 | 0.640 | 0.049 | 0.100 | 0.166 |
|  | Integrated | CoxPH | 0.652 ± 0.066 | 0.646 (0.571–0.716) | 0.682 | 0.660 | 0.586 | 0.647 | 0.047 | 0.099 | 0.157 |
|  |  | XGBoost | 0.682 ± 0.064 | 0.682 (0.612–0.754) | 0.749 | 0.634 | 0.675 | 0.733 | 0.049 | 0.101 | 0.156 |
|  |  | RSF | 0.685 ± 0.066 | 0.685 (0.622–0.751) | 0.767 | 0.615 | 0.655 | 0.737 | 0.048 | 0.099 | 0.153 |
| K = 3<sup>a</sup> | Clinical only | CoxPH | 0.648 ± 0.073 | 0.645 (0.567–0.721) | 0.693 | 0.617 | 0.543 | 0.637 | 0.047 | 0.099 | 0.157 |
|  |  | XGBoost | 0.646 ± 0.091 | 0.642 (0.562–0.728) | 0.715 | 0.576 | 0.585 | 0.697 | 0.049 | 0.105 | 0.154 |
|  |  | RSF | 0.650 ± 0.088 | 0.648 (0.569–0.732) | 0.719 | 0.573 | 0.604 | 0.703 | 0.048 | 0.099 | 0.149 |
|  | Multi-omics only | CoxPH | 0.589 ± 0.075 | 0.583 (0.512–0.656) | 0.631 | 0.559 | 0.644 | 0.598 | 0.048 | 0.097 | 0.165 |
|  |  | XGBoost | 0.604 ± 0.052 | 0.606 (0.537–0.676) | 0.636 | 0.557 | 0.656 | 0.610 | 0.049 | 0.098 | 0.178 |
|  |  | RSF | 0.620 ± 0.063 | 0.618 (0.554–0.683) | 0.655 | 0.568 | 0.688 | 0.637 | 0.048 | 0.096 | 0.165 |
|  | Integrated | CoxPH | 0.666 ± 0.063 | 0.658 (0.588–0.728) | 0.709 | 0.634 | 0.628 | 0.671 | 0.048 | 0.098 | 0.159 |
|  |  | XGBoost | 0.677 ± 0.061 | 0.678 (0.605–0.750) | 0.725 | 0.624 | 0.654 | 0.709 | 0.049 | 0.103 | 0.171 |
|  |  | RSF | 0.691 ± 0.061 | 0.685 (0.620–0.754) | 0.748 | 0.634 | 0.668 | 0.733 | 0.047 | 0.097 | 0.151 |
| K = 5 | Clinical only | CoxPH | 0.660 ± 0.077 | 0.656 (0.576–0.735) | 0.701 | 0.615 | 0.571 | 0.665 | 0.047 | 0.099 | 0.154 |
|  |  | XGBoost | 0.663 ± 0.094 | 0.659 (0.574–0.745) | 0.716 | 0.624 | 0.576 | 0.702 | 0.047 | 0.101 | 0.147 |
|  |  | RSF | 0.660 ± 0.094 | 0.658 (0.574–0.750) | 0.716 | 0.595 | 0.602 | 0.703 | 0.047 | 0.097 | 0.148 |
|  | Multi-omics only | CoxPH | 0.596 ± 0.088 | 0.591 (0.521–0.667) | 0.628 | 0.582 | 0.648 | 0.622 | 0.048 | 0.096 | 0.164 |
|  |  | XGBoost | 0.601 ± 0.064 | 0.604 (0.533–0.670) | 0.618 | 0.582 | 0.655 | 0.631 | 0.049 | 0.098 | 0.177 |
|  |  | RSF | 0.599 ± 0.061 | 0.597 (0.527–0.663) | 0.620 | 0.585 | 0.655 | 0.623 | 0.049 | 0.098 | 0.167 |
|  | Integrated | CoxPH | 0.682 ± 0.075 | 0.676 (0.604–0.752) | 0.726 | 0.651 | 0.649 | 0.708 | 0.047 | 0.099 | 0.158 |
|  |  | XGBoost | 0.703 ± 0.064 | 0.711 (0.632–0.786) | 0.746 | 0.667 | 0.689 | 0.755 | 0.048 | 0.100 | 0.157 |
|  |  | RSF | 0.684 ± 0.073 | 0.681 (0.611–0.755) | 0.739 | 0.638 | 0.650 | 0.715 | 0.047 | 0.097 | 0.152 |
| Marginal Kaplan–Meier (null) |  |  | — | 0.500 | 0.500 | 0.500 | 0.500 | 0.500 | 0.047 | 0.099 | 0.168 |

<sup>a</sup> $K = 3$ is the primary analysis reported in the main manuscript (Table 2).
$K$ denotes the number of Sparse PCA components retained per modality. CV C-index is the mean ± SD across the 25 cross-validation folds. Pooled C-index and 95% confidence intervals were computed on pooled out-of-fold predictions using 1,000 bootstrap resamples. Uno's C is an inverse-probability-of-censoring-weighted estimator. Time-dependent AUC is based on 24, 43 and 64 deaths at 2, 3 and 5 years respectively; one-year values are not reported because only 11 deaths occurred within 12 months across the cohort. Brier scores should be read against the marginal Kaplan–Meier reference in the final row; lower is better.
Feature counts were 1, 4 and 5 at $K = 1$ ; 3, 12 and 15 at $K = 3$ ; and 5, 20 and 25 at $K = 5$ for the clinical-only, multi-omics-only and integrated feature sets respectively. At $K = 5$ the integrated feature set corresponds to 3.5 events per variable, below conventional recommendations.
RSF, random survival forest; IPCW, inverse probability of censoring weighting.

**Table S2:** Secondary analysis using a single stratified 70/30 partition (*K* = 3; training *n* = 412, test *n* = 177, 26 test events). Reported for comparison with the cross-validated estimates in Table 2 of the main manuscript.

| Feature set | Model | C-index (95% CI) | Time-dependent AUC |  |  | IPCW Brier |  |
| --- | --- | --- | --- | --- | --- | --- | --- |
|  |  |  | 2 y | 3 y | 5 y | 3 y | 5 y |
| Clinical only | CoxPH | 0.604 (0.448–0.756) | 0.564 | 0.460 | 0.627 | 0.125 | 0.168 |
|  | XGBoost | 0.542 (0.383–0.702) | 0.524 | 0.368 | 0.577 | 0.140 | 0.190 |
|  | RSF | 0.606 (0.467–0.741) | 0.646 | 0.408 | 0.581 | 0.131 | 0.172 |
| Multi-omics only | CoxPH | 0.627 (0.504–0.750) | 0.648 | 0.665 | 0.588 | 0.116 | 0.173 |
|  | XGBoost | 0.589 (0.456–0.742) | 0.504 | 0.665 | 0.587 | 0.115 | 0.199 |
|  | RSF | 0.604 (0.478–0.733) | 0.593 | 0.666 | 0.598 | 0.112 | 0.176 |
| Integrated | CoxPH | 0.657 (0.529–0.786) | 0.641 | 0.575 | 0.672 | 0.123 | 0.173 |
|  | XGBoost | 0.643 (0.513–0.783) | 0.588 | 0.584 | 0.689 | 0.125 | 0.187 |
|  | RSF | 0.669 (0.545–0.803) | 0.637 | 0.579 | 0.731 | 0.116 | 0.154 |

Time-dependent AUC in the test partition is based on 7, 15 and 20 deaths at 2, 3 and 5 years respectively, against 104, 77 and 43 event-free patients. Estimates at these event counts are imprecise and are reported only to illustrate the width of confidence intervals obtainable from a single partition, which are approximately twice those obtained by repeated cross-validation.
RSF, random survival forest; IPCW, inverse probability of censoring weighting.

**Table S3:**
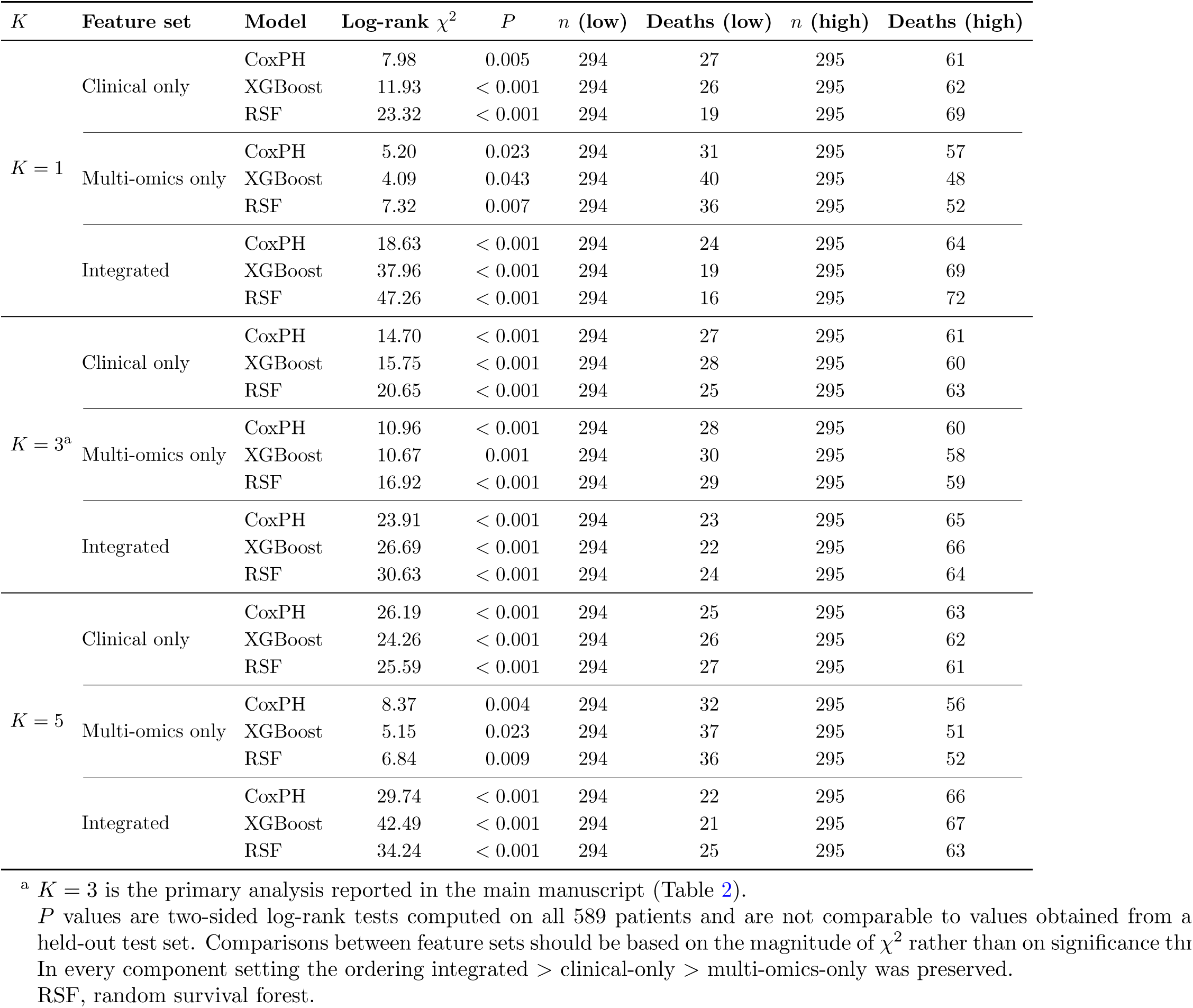
Kaplan–Meier risk stratification and log-rank test results across Sparse PCA component settings. Patients were dichotomised at the median pooled out-of-fold risk score (*n* = 589, 88 events).

| $K$ | Feature set | Model | Log-rank $\chi^2$ | $P$ | $n$ (low) | Deaths (low) | $n$ (high) | Deaths (high) |
| --- | --- | --- | --- | --- | --- | --- | --- | --- |
| $K = 1$ | Clinical only | CoxPH | 7.98 | 0.005 | 294 | 27 | 295 | 61 |
|  |  | XGBoost | 11.93 | < 0.001 | 294 | 26 | 295 | 62 |
|  |  | RSF | 23.32 | < 0.001 | 294 | 19 | 295 | 69 |
|  | Multi-omics only | CoxPH | 5.20 | 0.023 | 294 | 31 | 295 | 57 |
|  |  | XGBoost | 4.09 | 0.043 | 294 | 40 | 295 | 48 |
|  |  | RSF | 7.32 | 0.007 | 294 | 36 | 295 | 52 |
|  | Integrated | CoxPH | 18.63 | < 0.001 | 294 | 24 | 295 | 64 |
|  |  | XGBoost | 37.96 | < 0.001 | 294 | 19 | 295 | 69 |
|  |  | RSF | 47.26 | < 0.001 | 294 | 16 | 295 | 72 |
| $K = 3^a$ | Clinical only | CoxPH | 14.70 | < 0.001 | 294 | 27 | 295 | 61 |
|  |  | XGBoost | 15.75 | < 0.001 | 294 | 28 | 295 | 60 |
|  |  | RSF | 20.65 | < 0.001 | 294 | 25 | 295 | 63 |
|  | Multi-omics only | CoxPH | 10.96 | < 0.001 | 294 | 28 | 295 | 60 |
|  |  | XGBoost | 10.67 | 0.001 | 294 | 30 | 295 | 58 |
|  |  | RSF | 16.92 | < 0.001 | 294 | 29 | 295 | 59 |
|  | Integrated | CoxPH | 23.91 | < 0.001 | 294 | 23 | 295 | 65 |
|  |  | XGBoost | 26.69 | < 0.001 | 294 | 22 | 295 | 66 |
|  |  | RSF | 30.63 | < 0.001 | 294 | 24 | 295 | 64 |
| $K = 5$ | Clinical only | CoxPH | 26.19 | < 0.001 | 294 | 25 | 295 | 63 |
|  |  | XGBoost | 24.26 | < 0.001 | 294 | 26 | 295 | 62 |
|  |  | RSF | 25.59 | < 0.001 | 294 | 27 | 295 | 61 |
|  | Multi-omics only | CoxPH | 8.37 | 0.004 | 294 | 32 | 295 | 56 |
|  |  | XGBoost | 5.15 | 0.023 | 294 | 37 | 295 | 51 |
|  |  | RSF | 6.84 | 0.009 | 294 | 36 | 295 | 52 |
|  | Integrated | CoxPH | 29.74 | < 0.001 | 294 | 22 | 295 | 66 |
|  |  | XGBoost | 42.49 | < 0.001 | 294 | 21 | 295 | 67 |
|  |  | RSF | 34.24 | < 0.001 | 294 | 25 | 295 | 63 |

<sup>a</sup> $K = 3$ is the primary analysis reported in the main manuscript (Table 2).
$P$ values are two-sided log-rank tests computed on all 589 patients and are not comparable to values obtained from a held-out test set. Comparisons between feature sets should be based on the magnitude of $\chi^2$ rather than on significance thresholds. In every component setting the ordering integrated > clinical-only > multi-omics-only was preserved. RSF, random survival forest.

**Table S4:** Conventional Cox proportional hazards models fitted to structured clinical variables. Panel A gives hazard ratios for the primary baseline model; Panel B gives cross-validated performance for three nested specifications, using folds, seed and stratification identical to the main analysis (*n* = 589, 88 events).

**Panel A. Hazard ratios, primary baseline model (age + stage)**
| Variable | Hazard ratio | 95% CI | $P$ |
| --- | --- | --- | --- |
| Age (per SD) | 1.47 | 1.19–1.81 | < 0.001 |
| Stage II vs Stage I | 1.18 | 0.67–2.09 | 0.573 |
| Stage III vs Stage I | 1.62 | 0.85–3.08 | 0.143 |

**Panel B. Cross-validated performance**
| Model | $p$ | CV C-index | Pooled C | Uno's | Time-dependent AUC | | | IPCW Brier | |
| --- | --- | --- | --- | --- | --- | --- | --- | --- | --- |
| | | (mean ± SD) | (95% CI) | C | 2 y | 3 y | 5 y | 3 y | 5 y |
| Marginal Kaplan–Meier | 0 | 0.500 | 0.500 | 0.500 | 0.500 | 0.500 | 0.500 | 0.099 | 0.166 |
| Age + stage<sup>a</sup> | 3 | 0.636 ± 0.053 | 0.637 (0.559–0.707) | 0.549 | 0.697 | 0.642 | 0.610 | 0.095 | 0.162 |
| Age + stage + race<sup>b</sup> | 5 | 0.620 ± 0.069 | 0.616 (0.532–0.693) | 0.532 | 0.675 | 0.597 | 0.594 | 0.096 | 0.163 |
| Full clinical<sup>c</sup> | 9 | 0.680 ± 0.088 | 0.679 (0.603–0.760) | 0.721 | 0.656 | 0.596 | 0.693 | 0.101 | 0.149 |

**Panel C. Kaplan–Meier stratification at the median risk score**
| Model | Log-rank $\chi^2$ | $P$ | Hazard ratio (95% CI) | Deaths, low / high |
| --- | --- | --- | --- | --- |
| Age + stage | 6.30 | 0.012 | 1.70 (1.12–2.60) | 40/294 vs 48/295 |
| Age + stage + race | 6.85 | 0.009 | 1.74 (1.14–2.66) | 39/294 vs 49/295 |
| Full clinical | 23.65 | < 0.001 | 3.02 (1.89–4.83) | 24/294 vs 64/295 |

<sup>a</sup> Primary clinical reference model, reported in Table 2 of the main manuscript.
<sup>b</sup> Adding race did not improve model fit (likelihood ratio $\chi^2 = 1.09$ , $df = 2$ , $P = 0.579$ ) and slightly reduced cross-validated discrimination.
<sup>c</sup> Additionally includes adjuvant therapy (likelihood ratio $\chi^2 = 69.78$ , $df = 4$ , $P < 0.001$ versus the preceding model). Therapy is assigned after diagnosis and is influenced by prognosis; the apparent effect (hazard ratio 0.18, 95% CI 0.08–0.40 for combined therapy versus none, and 0.20, 0.09–0.47 for pharmaceutical therapy alone) most plausibly reflects confounding by indication rather than prognostic information available at diagnosis. This model is reported as an information-preservation comparator for the BioBERT encoding of the same variables, not as a prognostic baseline.
$p$ , number of covariates. Pathologic stage was collapsed to Stage I–IV; Stage IV ( $n = 9$ ) and Unknown ( $n = 1$ ) were merged into the reference category because fewer than ten patients were available. Confidence intervals are from 1,000 bootstrap resamples.

**Figure S1:**
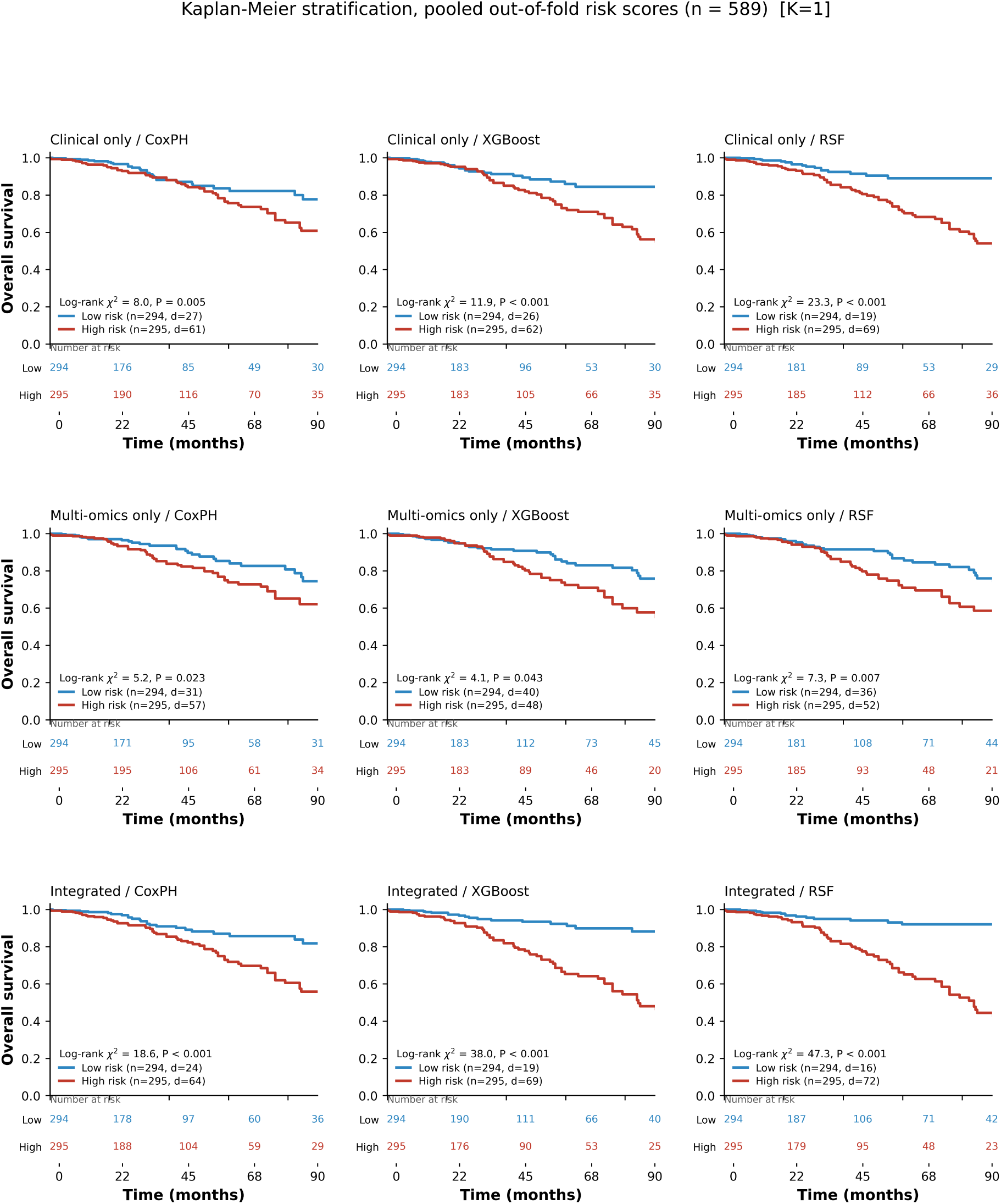
Kaplan–Meier risk stratification for CoxPH, XGBoost and random survival forest models using clinical-only, multi-omics-only and integrated feature sets, with *K* = 1 Sparse PCA component per modality. Patients were dichotomised at the median pooled out-of-fold risk score. Number-at-risk tables appear beneath each panel.

**Figure S2:**
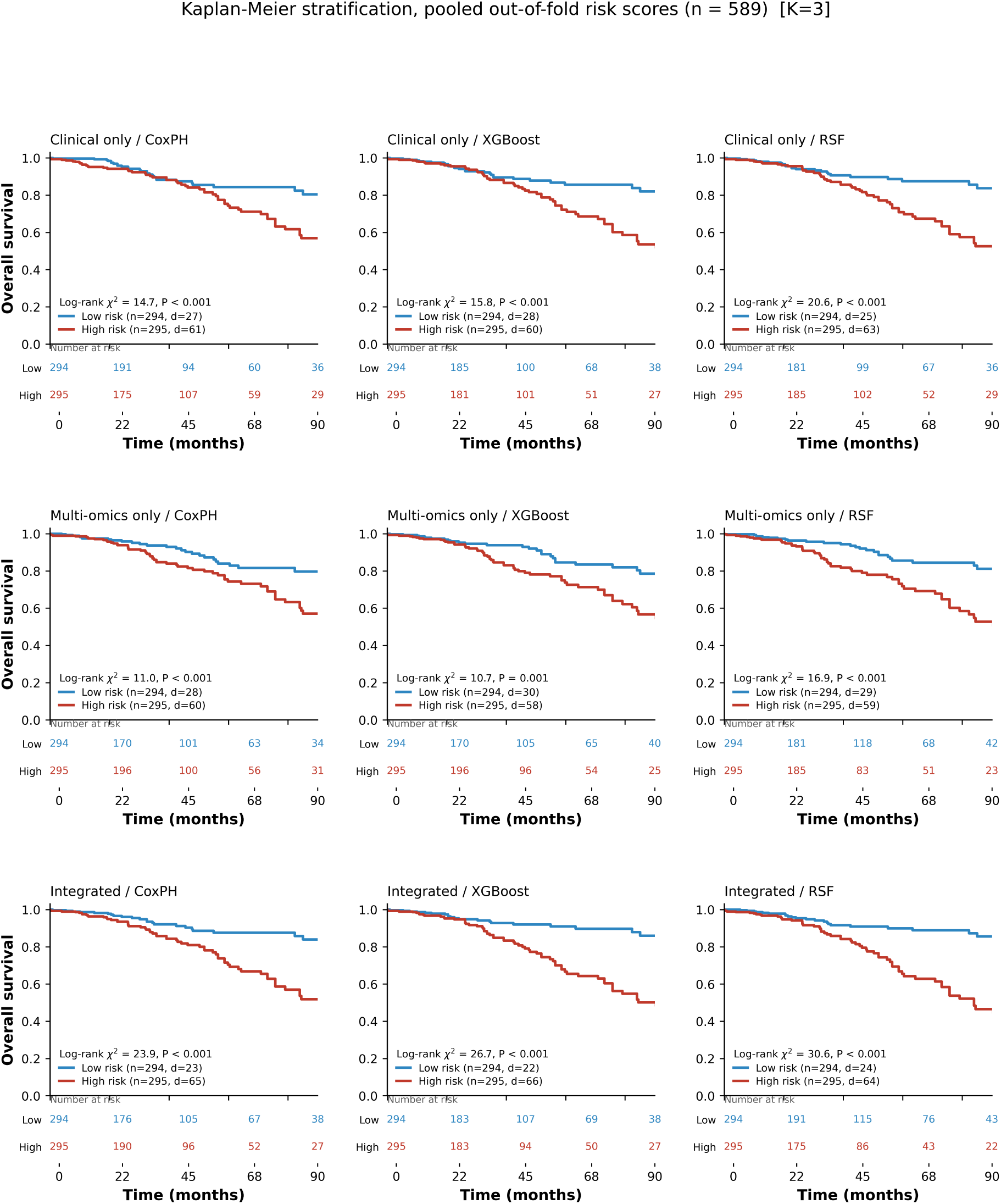
Kaplan–Meier risk stratification for all feature sets and algorithms with *K* = 3 Sparse PCA components per modality (primary analysis).

**Figure S3:**
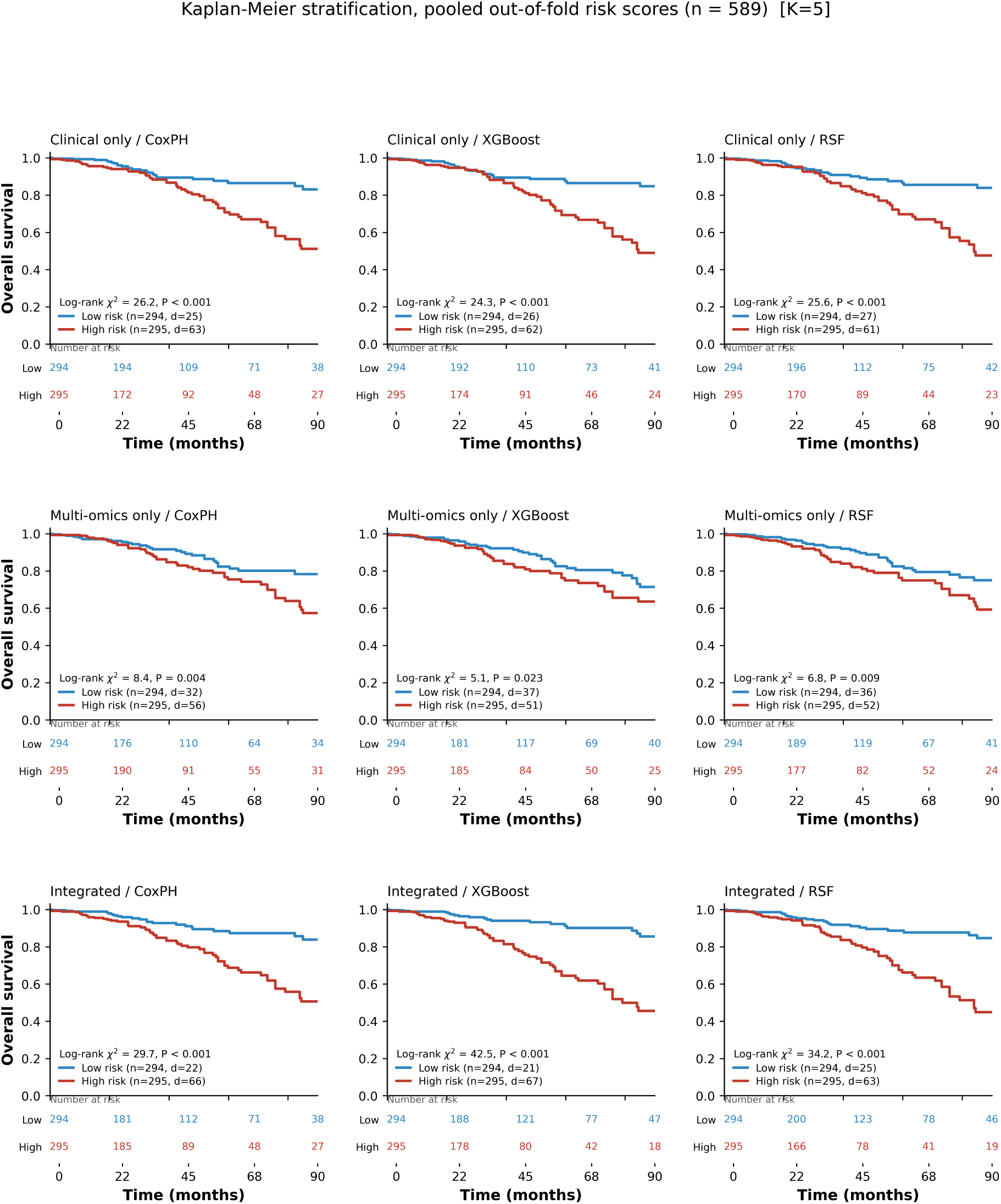
Kaplan–Meier risk stratification for all feature sets and algorithms with *K* = 5 Sparse PCA components per modality.

**Figure S4:**
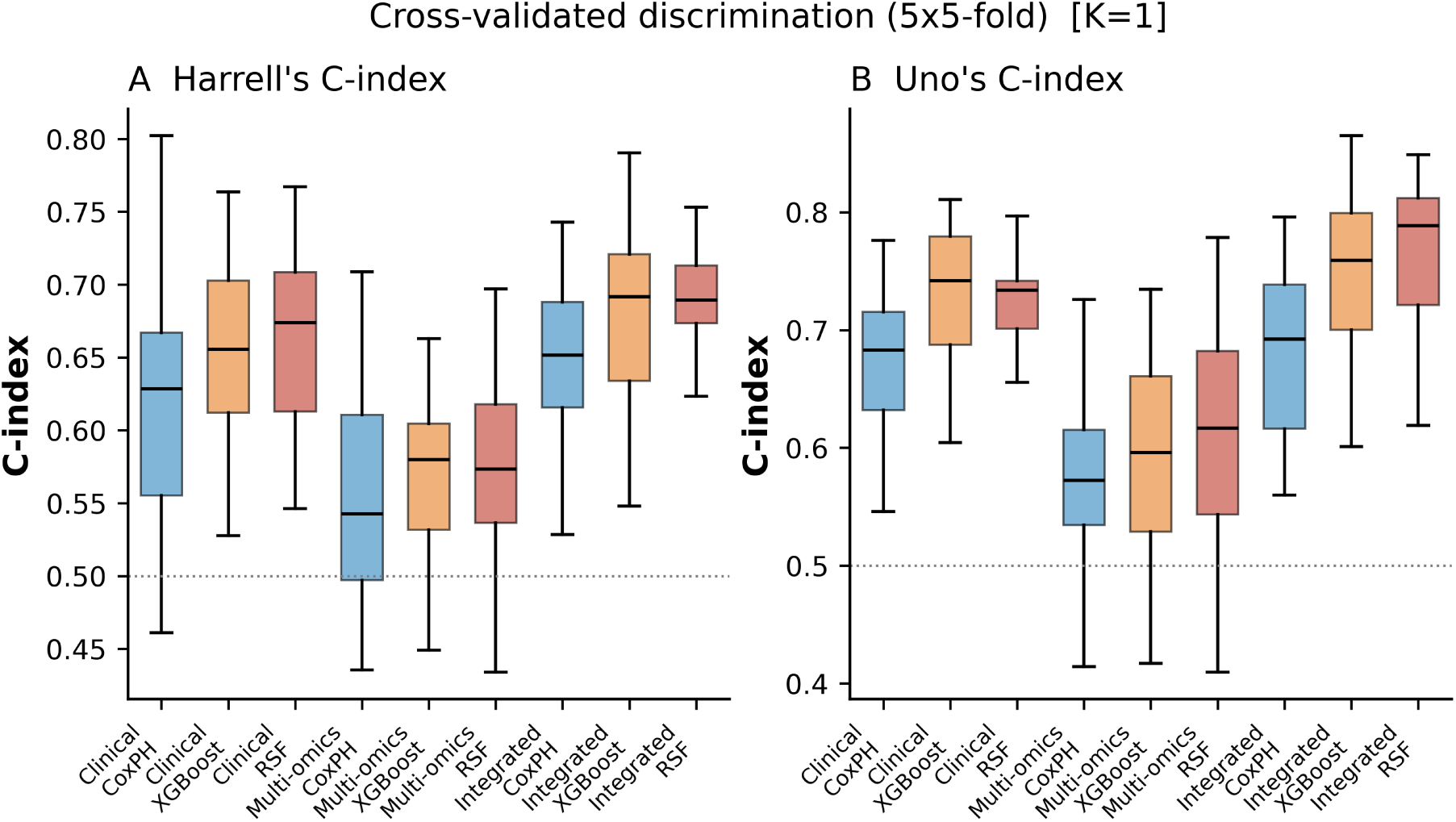
Distribution of Harrell’s and Uno’s concordance indices across the 25 cross-validation folds, *K* = 1. Boxes show the interquartile range; points are individual folds.

**Figure S5:**
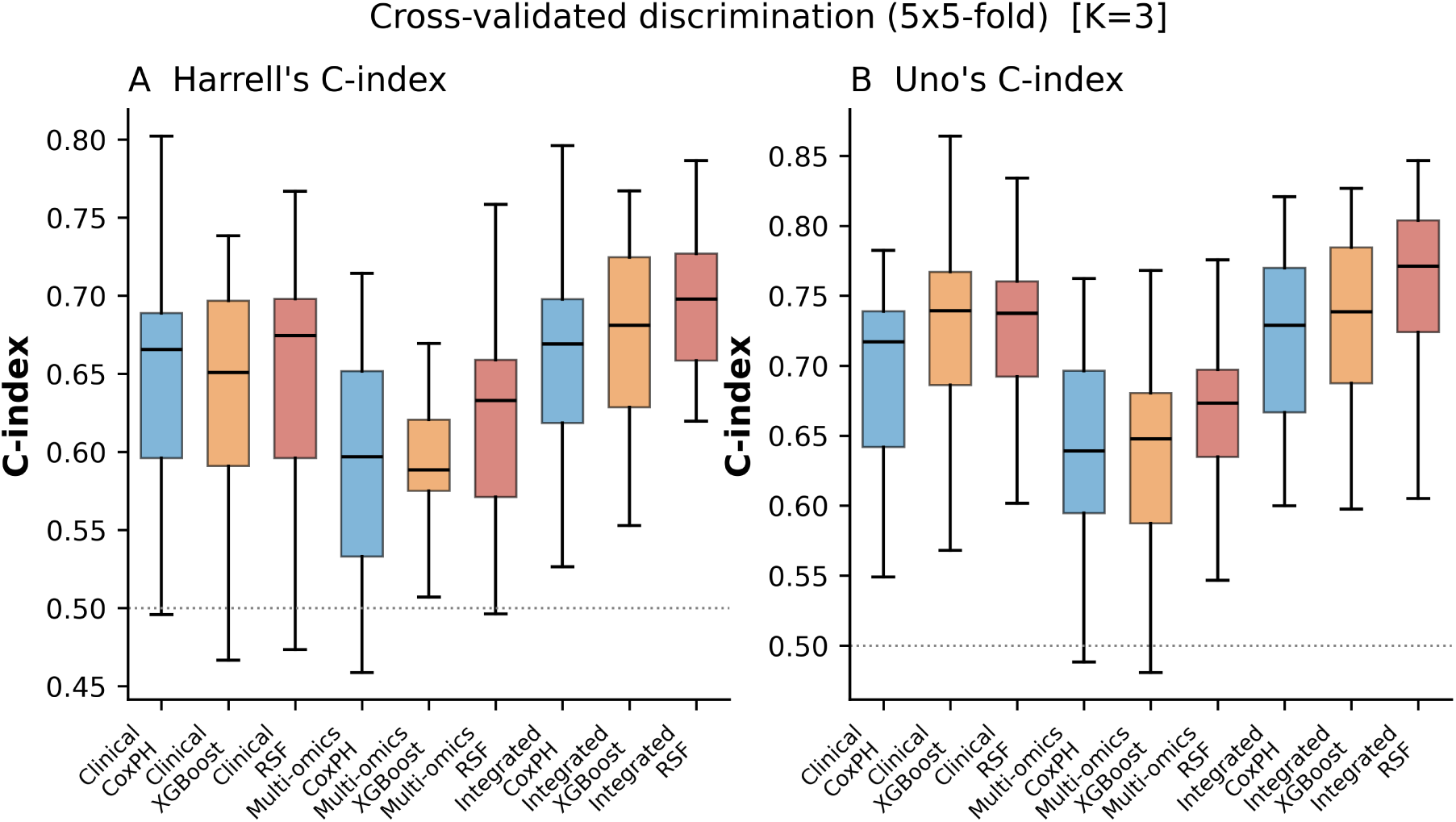
Distribution of Harrell’s and Uno’s concordance indices across the 25 cross-validation folds, *K* = 3 (primary analysis). The spread of these distributions illustrates the precision available at 88 events.

**Figure S6:**
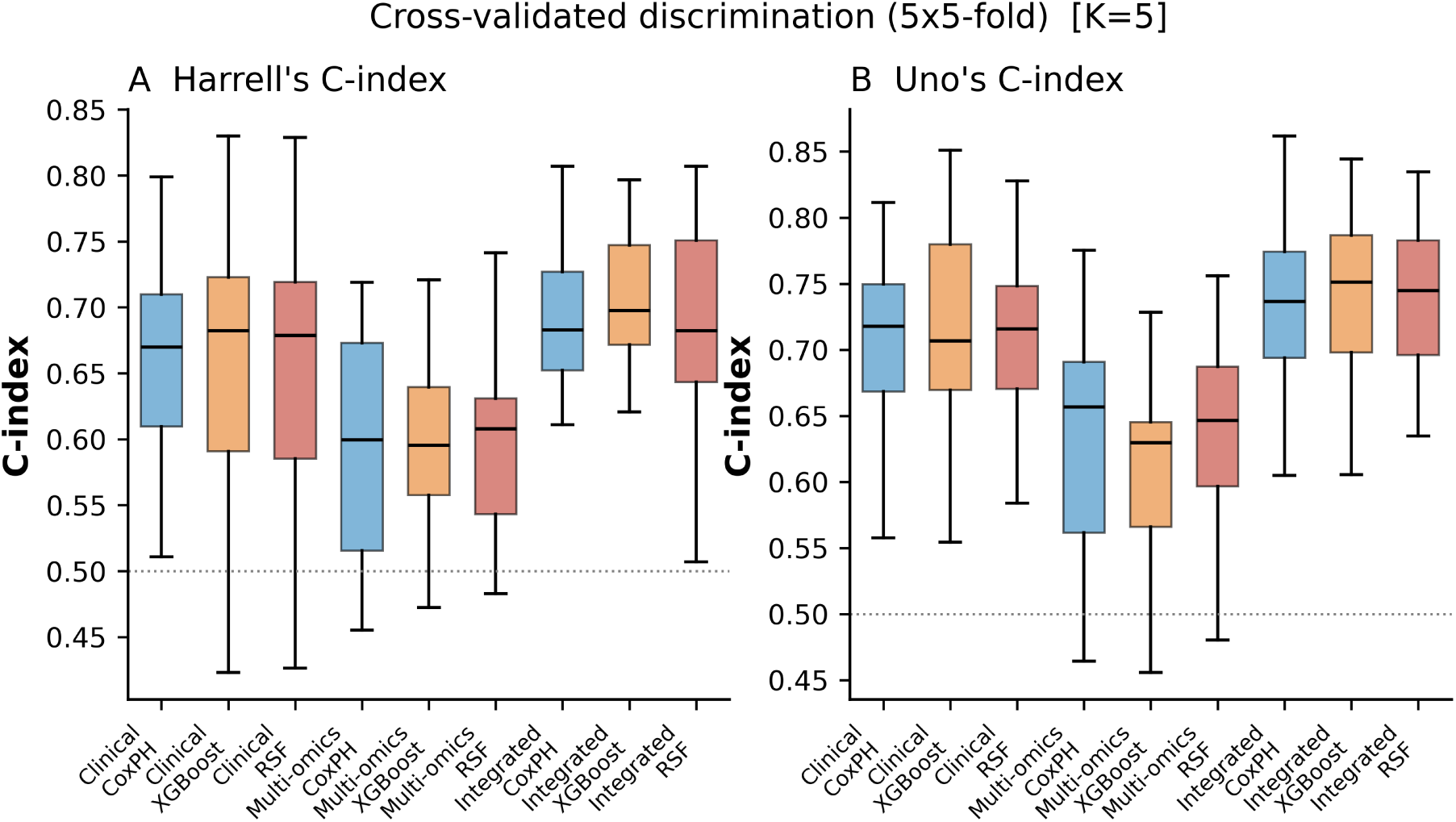
Distribution of Harrell’s and Uno’s concordance indices across the 25 cross-validation folds, *K* = 5.

**Figure S7:**
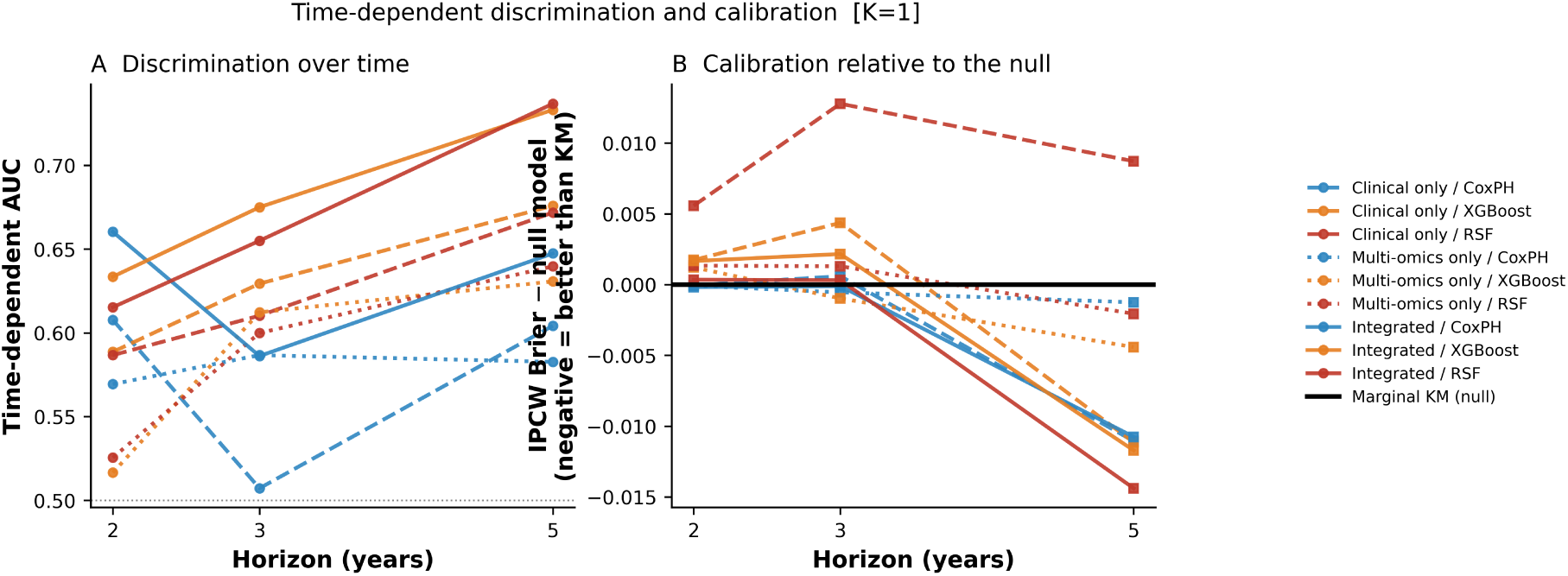
Time-dependent AUC (A) and IPCW Brier score relative to a marginal Kaplan–Meier reference (B), *K* = 1. Values below zero in panel B indicate better calibration than the population survival curve.

**Figure S8:**
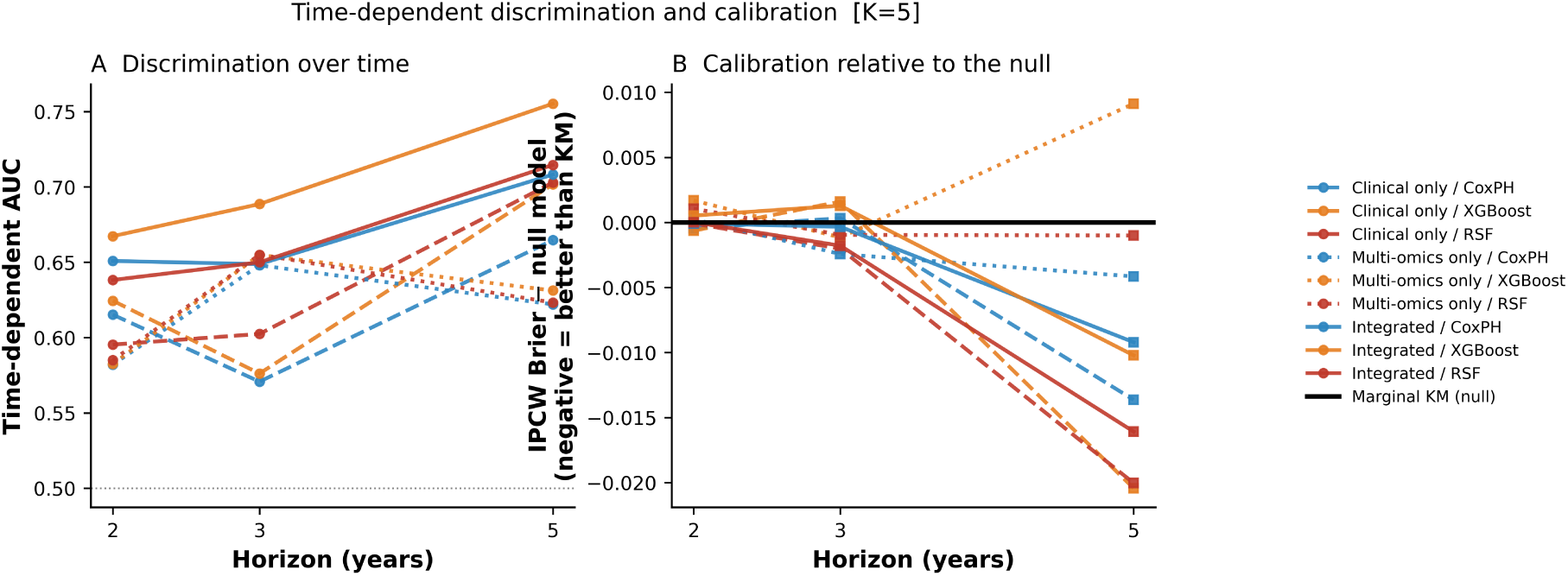
Time-dependent AUC (A) and IPCW Brier score relative to a marginal Kaplan–Meier reference (B), *K* = 5.

**Figure S9:**
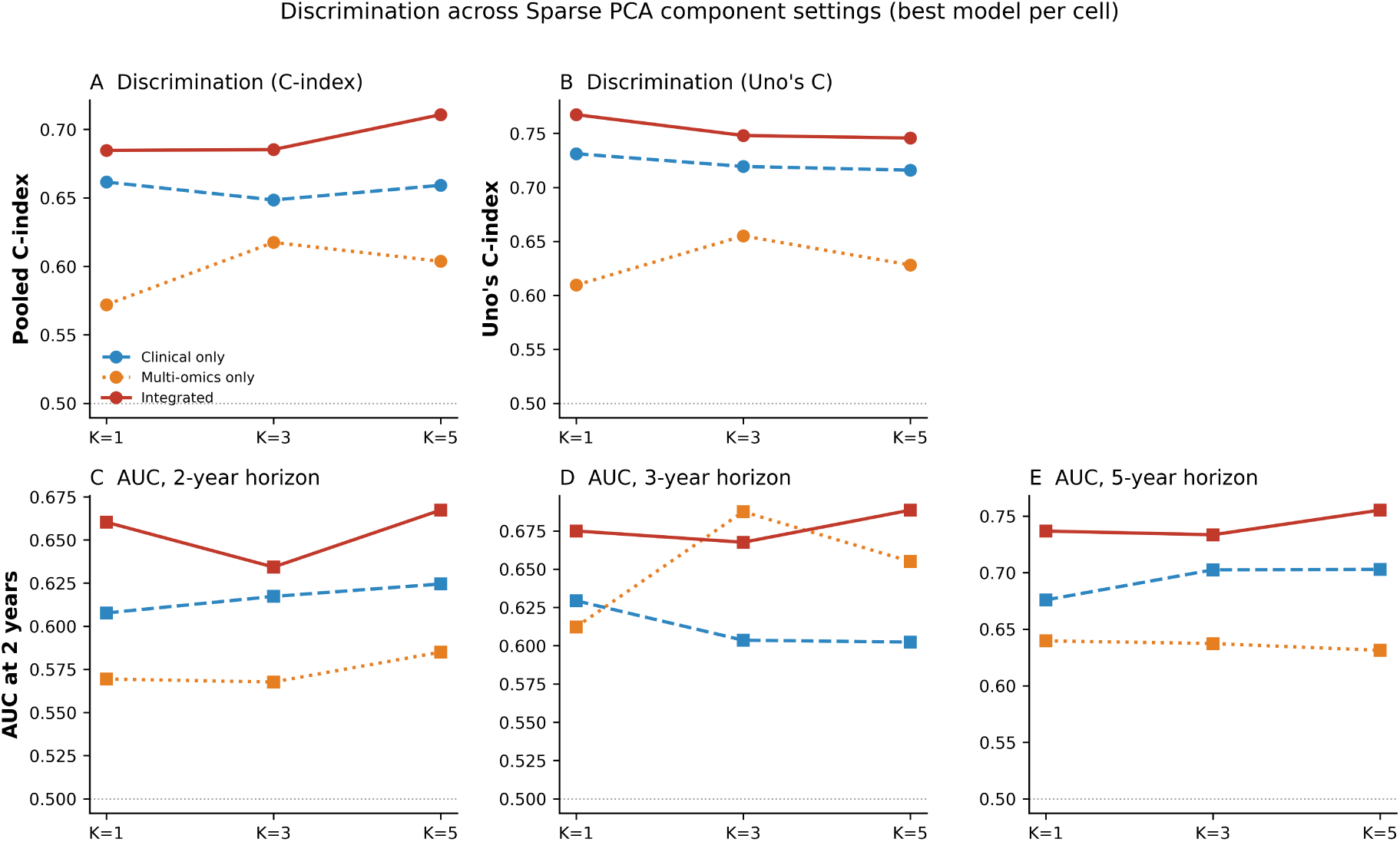
Concordance and time-dependent AUC across Sparse PCA component settings (*K* = 1, 3 and 5) for each feature set. Integrated discrimination was essentially unchanged across settings, whereas multi-omics-only discrimination peaked at *K* = 3.

